# Characteristics and Determinants of Pulmonary Long COVID

**DOI:** 10.1101/2024.02.13.24302781

**Authors:** Michael John Patton, Donald Benson, Sarah W. Robison, Raval Dhaval, Morgan L. Locy, Kinner Patel, Scott Grumley, Emily B. Levitan, Peter Morris, Matthew Might, Amit Gaggar, Nathaniel Erdmann

## Abstract

**RATIONALE:** Persistent cough and dyspnea are prominent features of post-acute sequelae of SARS-CoV-2 (termed ’Long COVID’); however, physiologic measures and clinical features associated with these pulmonary symptoms remain poorly defined.

**OBJECTIVES:** Using longitudinal pulmonary function testing (PFTs) and CT imaging, this study aimed to identify the characteristics and determinants of pulmonary Long COVID.

**METHODS:** The University of Alabama at Birmingham Pulmonary Long COVID cohort was utilized to characterize lung defects in patients with persistent pulmonary symptoms after resolution primary COVID infection. Longitudinal PFTs including total lung capacity (TLC) and diffusion limitation of carbon monoxide (DLCO) were used to evaluate restriction and diffusion impairment over time in this cohort. Analysis of chest CT imaging was used to phenotype the pulmonary Long COVID pathology. Risk factors linked to development of pulmonary Long COVID were estimated using univariate and multivariate logistic regression models.

**MEASUREMENTS AND MAIN RESULTS:** Longitudinal evaluation 929 patients with post-COVID pulmonary symptoms revealed diffusion impairment (DLCO ≤80%) and restriction (TLC ≤80%) in 51% of the cohort (n=479). In multivariable logistic regression analysis (adjusted odds ratio; aOR, 95% confidence interval [CI]), invasive mechanical ventilation during primary infection conferred the greatest increased odds of developing pulmonary Long COVID with diffusion impaired restriction (aOR=10.9 [4.09-28.6]). Finally, a sub-analysis of CT imaging identified evidence of fibrosis in this population.

**CONCLUSIONS:** Persistent diffusion impaired restriction was identified as a key feature of pulmonary Long COVID. Subsequent clinical trials should leverage combined symptomatic and quantitative PFT measurements for more targeted enrollment of pulmonary Long COVID patients.

## INTRODUCTION

A major consequence of the COVID-19 pandemic has been the complex and frequently debilitating post-acute sequelae of SARS-CoV-2 infection (PASC, also termed ’Long COVID’), estimated to occur in 10% of patients after primary infection.^1,2^ To date, over 150 distinct Long COVID symptoms involving every major organ system have been reported.^3^ Recent efforts to develop a consensus definition for Long COVID using symptom clustering have highlighted broad disease sub-types associated with chronic fatigue, post-exertional malaise, brain fog, and loss of smell or taste.^4^ These efforts represent an important first step for Long COVID research; however, characterizing discreet endotypes with widely available quantitative physiologic measurements is essential for standardizing Long COVID diagnosis and management.

Prolonged pulmonary symptoms, notably dyspnea and cough, are among the most commonly reported manifestations of Long COVID (hereon referred to as ’pulmonary Long COVID’).^4–6^ Although pulmonary symptoms are frequently reported, the underlying cause(s) and clinical trajectory of Long COVID patients suffering from these symptoms remains unclear. Prior studies have suggested diffusion impairment that resolves within 1-year of hospitalization is a common feature of post-acute COVID-19.^7–10^ Other post-acute COVID follow-up studies have reported radiologic evidence of lung pathology characterized by ground glass opacities and fibrotic changes.^11–18^ These results have provided valuable insight into post-COVID lung pathology, but are limited by small cohort size and inherently biased towards post-acute COVID patients, rather than Long COVID populations presenting for persistent pulmonary complaints.

To address this knowledge gap, we leveraged a large, demographically diverse cohort exclusively comprised of Long COVID patients experiencing pulmonary symptoms persisting for ≥1 month after resolution of acute SARS-CoV-2 infection. Using longitudinal pulmonary function testing (PFT) and computerized tomography (CT) imaging, we identify specific disease features and risk factors linked to the development of pulmonary Long COVID. These presentations capture the population of patients with persistent clinical symptoms and reflect the natural history of pulmonary Long COVID. Lastly, we provide evidence for a novel endotype of Long COVID defined by persistent diffusion impairment and pulmonary restriction.

## METHODS

### Study Design and Population

This single-center, retrospective cohort study was performed among adult patients (≥18 years) with a positive COVID-19 PCR and/or rapid antigen test during the study window (03/2020-08/2023), followed by self- or physician referral to the University of Alabama at Birmingham (UAB) Post-COVID Pulmonary Clinic for complaint of unresolved respiratory symptoms. Pulmonary Long COVID was defined as pulmonary symptoms persisting for ≥1 month that had developed ≥28 days after resolution of primary SARS-CoV-2 infection. Baseline (1st visit) PFTs and CT scans were defined by the first date of the measurement within a window of 14-days prior to and 6-months after the patient’s 1st Long COVID clinic visit date. Follow-up measurements (termed 2nd and 3rd visit) were restricted to the study window and had to occur after the first encounter measurement. The cohort was stratified by percent predicted diffusion capacity for carbon monoxide (DLCO; normal=DLCO >80%, impaired=DLCO ≤80%) and percent predicted total lung capacity (TLC) in accord with the American Thoracic Society consensus definition for restrictive lung disease. Lung restriction sub-groupings were (1) no restriction TLC >80%, (2) mild restriction TLC 71-80%, (3) moderate restriction TLC 51-70%, (4) severe restriction TLC ≤50%.^19,20^ All pulmonary function studies were conducted utilizing the same equipment (Vyntus^TM^ system from Vyaire Medical Incorporated).

### Patient Variables Extracted from Electronic Medical Record and Radiographic Images

Clinical variables were extracted for all patients in the cohort including: advanced age (≥65 years), biological sex, elevated BMI (≥30), smoking history (never, former, or current smoker), pre-COVID vaccination status, ICU admission, COVID-19 severity (assessed using the WHO score system representing maximum oxygen therapy support required), and therapeutics (dexamethasone, remdesivir, days on a specific oxygen support device) during primary infection, SARS-CoV-2 variant, and months from primary infection to first Long COVID clinic visit.^21^ Pre-COVID comorbidities (renal, pulmonary, heart failure or hypertension, diabetes, obstructive sleep apnea, and immunosuppression) were determined by physician review of medical records. Chest CT imaging was assessed by two blinded cardiothoracic radiologists for the presence or absence of 6 pulmonary imaging findings: lung consolidation, ground-glass opacities, reticulations, other fibrotic-like changes (i.e. architectural distortion, traction bronchiectasis and honeycombing; termed ‘other fibrosis’), bronchiectasis, and emphysema (see eTable 3 for two-reader similarity evaluation). An overall severity score was determined using a previously defined image CT scoring system quantifying abnormalities in all 5 lung lobes with scores ranging from 0 (no involvement) to 25 (multi-lobe involvement).^22^ Detailed information on variables extracted can be found in the supplemental materials (eTable 1).

### Statistical Analysis

Cohort statistics were reported with mean ±standard deviation or median [Q1-Q3] in Table 1. Alluvial diagrams were used to assess relative DLCO and TLC improvement or decline in patients with 3-consecutive follow-ups visits (Figure 2C). Logistic regression models (unadjusted variable and multi-variable adjusted) were used to discover risk factors for developing diffusion impairment (DLCO≤80%) with severe or moderate restriction (TLC≤70%) at 1st Long COVID clinic visit (Table 2; see eTable 1 for details on model variables and eTables 5-7 for model sensitivity testing omitting patients without a primary COVID infection hospitalization and patients with pre-existing pulmonary comorbities). All modeling results are reported as either unadjusted (OR) or adjusted odds ratios (aOR) with bootstrapped (n=1000 iterations) 95% confidence intervals (CI). All statistical analyses were performed using R (version 4.2, R Foundation).

**Table 1:**
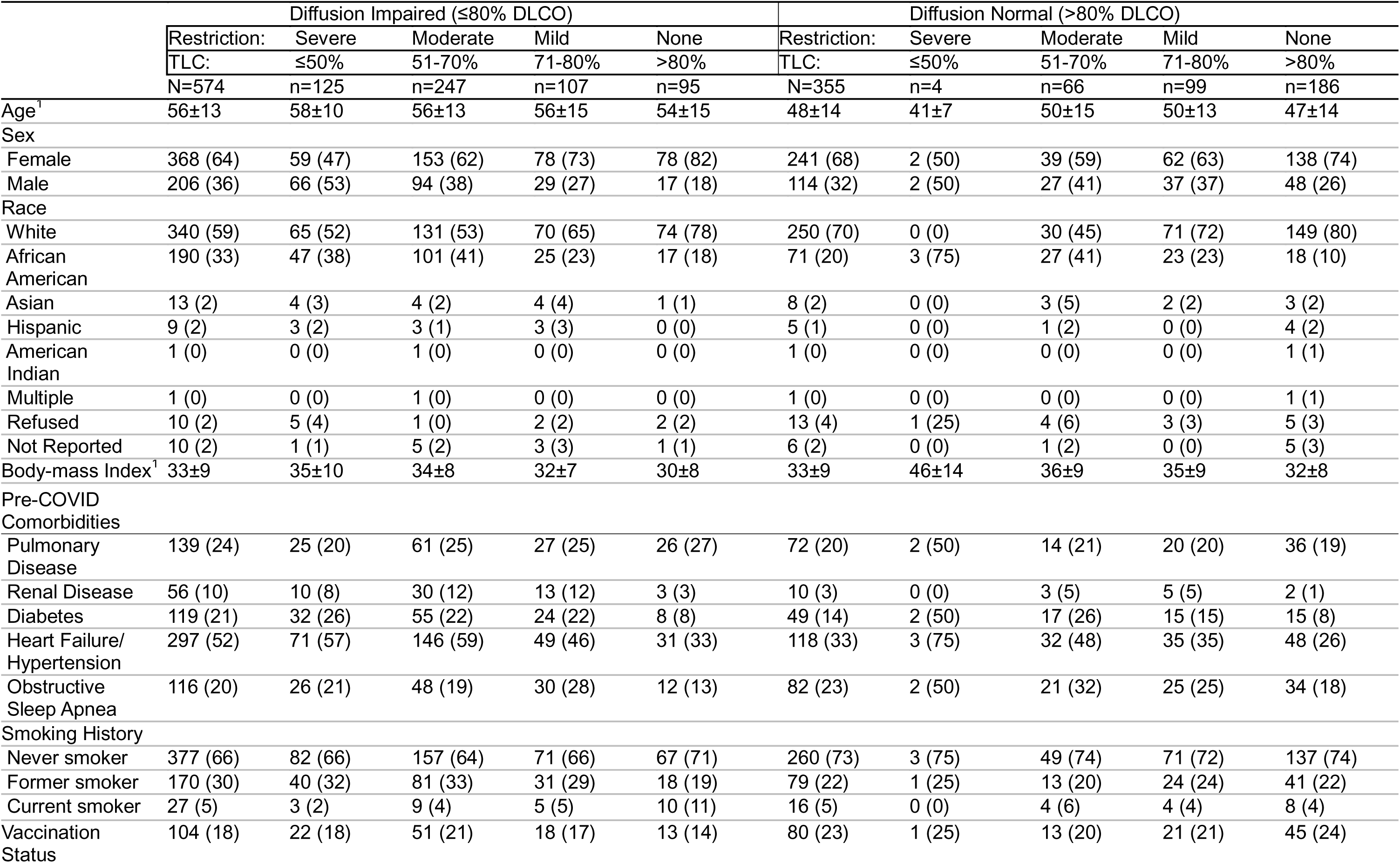
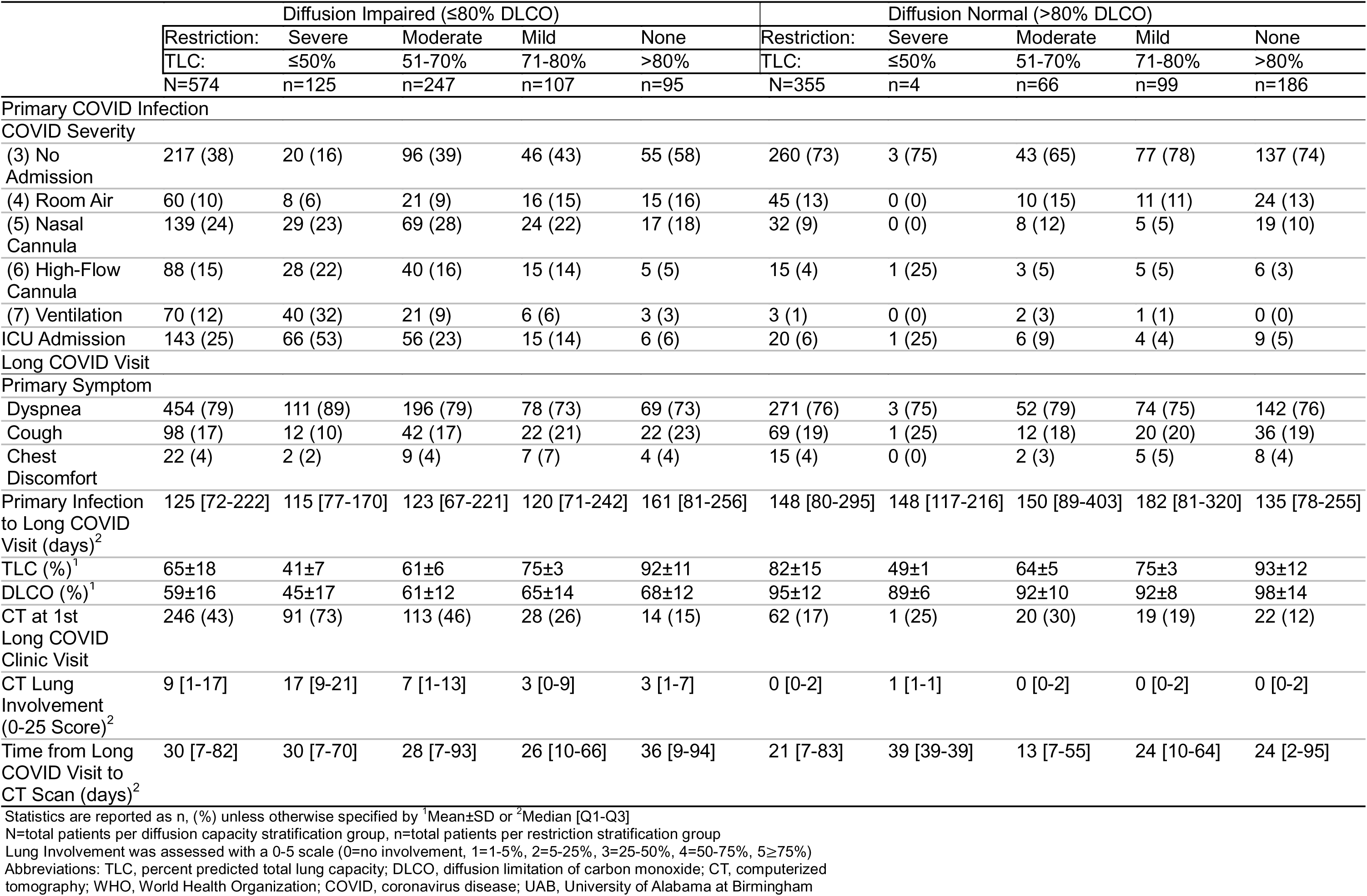
Characteristics of Pulmonary Long COVID Patients Stratified by 1^st^ Visit Diffusion Capacity and Restriction in the UAB Cohort.

**Table 2:** Risk Factors for Pulmonary Long COVID with Diffusion Impaired Restriction.

|  | Patients<br>Total N | Diffusion Impaired<br>Restriction<sup>2</sup><br>N (% of Total) | Diffusion Impaired<br>Restriction<br>Unadjusted OR [95% CI]<sup>1</sup> | Diffusion Impaired<br>Restriction<br>Adjusted OR [95% CI]<sup>1</sup> |
| --- | --- | --- | --- | --- |
| **Advanced Age** |  |  |  |  |
| <65 years | 743 | 280 (38) | — | — |
| ≥65 years | 186 | 92 (49) | 1.63 [1.23-2.23] | 1.22 [0.81-1.88] |
| **Sex** |  |  |  |  |
| Female | 609 | 212 (35) | — | — |
| Male | 320 | 160 (50) | 1.88 [1.43-2.48] | 1.44 [1.03-1.98] |
| **Elevated BMI** |  |  |  |  |
| <30 | 376 | 138 (37) | — | — |
| ≥30 | 553 | 234 (42) | 1.28 [0.97-1.66] | 1.27 [0.91-1.77] |
| **Pulmonary Disease** |  |  |  |  |
| No | 718 | 286 (40) | — | — |
| Yes | 211 | 86 (41) | 1.04 [0.76-1.41] | 0.95 [0.67-1.36] |
| **Renal Disease** |  |  |  |  |
| No | 863 | 332 (38) | — | — |
| Yes | 66 | 40 (61) | 2.48 [1.47-4.15] | 1.03 [0.57-1.99] |
| **Diabetes** |  |  |  |  |
| No | 761 | 285 (37) | — | — |
| Yes | 168 | 87 (52) | 1.82 [1.30-2.51] | 1.20 [0.79-1.78] |
| **Heart Failure or Hypertension** |  |  |  |  |
| No | 514 | 155 (30) | — | — |
| Yes | 415 | 217 (52) | 2.54 [1.98-3.32] | 2.09 [1.46-2.99] |
| **Obstructive Sleep Apnea** |  |  |  |  |
| No | 731 | 298 (41) | — | — |
| Yes | 198 | 74 (37) | 0.88 [0.63-1.17] | 0.51 [0.32-0.72] |
| **Smoking History** |  |  |  |  |
| Never smoker | 637 | 239 (38) | — | — |
| Current or Former Smoker | 292 | 133 (46) | 1.39 [1.03-1.86] | 1.25 [0.89-1.74] |
| **Vaccination Status** |  |  |  |  |
| No | 745 | 299 (40) | — | — |
| Yes | 184 | 73 (40) | 0.99 [0.72-1.39] | 1.34 [0.89-1.99] |
| **Months from Primary Infection**<br>**to Long COVID Clinic Visit** |  |  |  |  |
| 1-3 Months | 306 | 132 (43) | — | — |
| 3-6 Months | 292 | 139 (48) | 1.19 [0.88-1.64] | 0.96 [0.67-1.39] |
| 6-12 Months | 197 | 59 (30) | 0.56 [0.38-0.81] | 0.70 [0.44-1.10] |
| >12 Months | 134 | 42 (31) | 0.61 [0.40-0.89] | 0.64 [0.37-1.09] |
| **ICU Admission** |  |  |  |  |
| No | 766 | 250 (33) | — | — |
| Yes | 163 | 122 (75) | 6.15 [4.24-9.36] | 1.48 [0.71-3.27] |
| **COVID Severity (WHO Score)** |  |  |  |  |
| (3) No Admission | 477 | 116 (24) | — | — |
| (4) Room Air | 105 | 29 (28) | 1.20 [0.71-1.93] | 1.03 [0.57-1.73] |
| (5) Nasal Cannula | 171 | 98 (57) | 4.21 [2.90-6.07] | 4.00 [2.61-6.33] |
| (6) High-Flow Cannula | 103 | 68 (66) | 6.10 [3.92-9.82] | 3.86 [1.73-8.09] |
| (7) Ventilation | 73 | 61 (84) | 16.0 [8.87-34.4] | 10.9 [4.09-28.6] |

<sup>1</sup>Unadjusted odds ratio (OR) and adjusted odds ratio (aOR), 95% confidence interval (n=1000 bootstraps)<sup>2</sup>Diffusion impaired restriction is defined by a DLCO ≤80% and a TLC ≤70% measured by PFT at the 1<sup>st</sup> Long COVID clinic visit

## RESULTS

### Study Population

A total of 1,097 patients with prolonged pulmonary symptoms after primary SARSCoV-2 infection were identified based upon their evaluation in post-acute COVID pulmonary clinic. After exclusion, 929 patients with prolonged pulmonary symptoms (dyspnea, cough, or chest discomfort) lasting ≥1 month after resolution of primary SARS-CoV-2 infection and a subsequent PFT were included in this study (Figure 1, Table 1). To evaluate the role of pulmonary abnormalities in DLCO and evidence of pro-fibrotic processes, we stratified patients by diffusion impairment and severity of pulmonary restriction as measured by first post-acute PFT. Median time from primary SARS-CoV-2 infection to 1st Long COVID clinic visit was 125 days for diffusion impaired patients and 148 days for patients with normal diffusion capacity (Table 1). Patient age (mean, ±SD) for the diffusion impaired group was 56±13 years compared to 48±14 years in the normal diffusion group (Table 1). For both groups, the majority of primary SARS-CoV-2 infections occurred during the alpha-variant wave (range: 62-63%), followed by 21-23% and 15-16% occurring in the delta and omicron waves, respectively (eTable 2). Differences in primary pulmonary symptom reported at 1st Long COVID clinic visit were unremarkable between patients with and without diffusion impairment (range; dyspnea: 76-79%, cough: 17-19%, chest discomfort: 4%).

**Figure 1:**
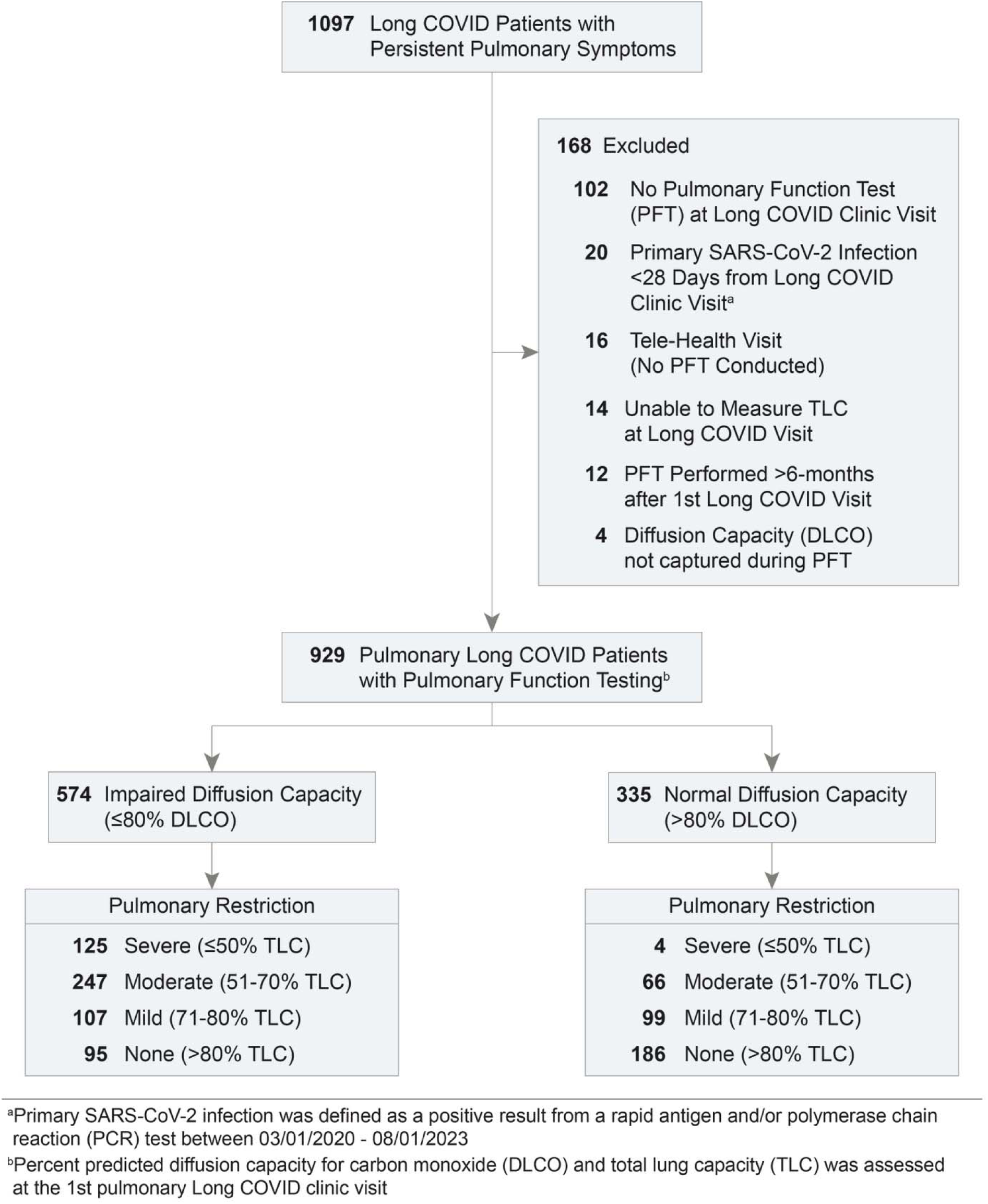
Accrual of Long COVID Patients with Persistent Pulmonary Symptoms.

We sought to determine how acute disease severity contributed to PFT findings for patients with pulmonary Long COVID. Acute disease severity was stratified by peak WHO ordinal score. Of the 73 patients who required invasive mechanical ventilation (IMV), only 3 were in the normal diffusion capacity group (Table 1). Increased frequency and duration of oxygen support was observed among diffusion impaired patients with the severe restriction group (IMV: 32%, HFNC: 22%) exceeding that of the moderate (IMV: 9%, HFNC: 16%), mild (IMV: 6%, HFNC: 14%), and no restriction groups (IMV: 3%, HFNC: 5%; Table 1). A similar trend of increased time of oxygen therapy (median [Q1-Q3]) during primary infection was also observed in diffusion impaired patients (IMV: 18 [8-35] days; HFNC: 5 [3-8] days, and nasal cannula: 7 [4-11] days; eTable 2). Differences in vaccination, pre-existing pulmonary disease or obstructive sleep apnea, and smoking history prior to primary SARS-CoV-2 infection were minimal between diffusion impaired and normal groups (Table 1). Patients with diffusion impairment had a greater proportion of pre-existing diabetes (21%) and heart failure or hypertension (52%) compared to the normal diffusion group with 14% and 33%, respectively (Table 1). Cumulative lung involvement (25 points total, 5 points per lobe; median [Q1-Q3]) was higher in the diffusion impaired group (9 [1-17]) compared to the normal group (0 [0-2]). The degree of lung involvement on CT imaging for diffusion impaired patients correlated with increasing severity of lung restriction (none: 3 [1-7], mild: 3 [0-9], moderate: 7 [1-13], severe: 17 [9-21]; Table 1). Further characteristics of the cohort are presented in Table 1 and eTable 2.

### Longitudinal Evaluation of Pulmonary Function Testing

To assess physiological differences between patients with pulmonary Long COVID, we evaluated longitudinal PFTs over three total Long COVID visits. After 1st visit stratification, patients without diffusion impairment had, on average, normal lung capacity during the visit 2 visits (TLC 1st: 82±15%, 2nd: 79±13%) with mild decline on 3rd visit (72±15%) compared to patients with diffusion impairment on respective follow-up visits (TLC 1st: 65±18%, 2nd: 65±15%, 3rd: 64±15%) (Figure 2A, eTable 4). Diffusion impaired patients with severe or moderate restriction experienced little to no improvement in TLC at 2nd (severe: 51±14%, moderate: 66±11%) or 3rd visit (severe: 55±14%, moderate: 65±12%) (Figure 2A, eTable 4). Overall, patients with normal diffusion capacity at 1st visit maintained normal or above normal DLCO at the follow-up visits (DLCO 1st: 95±12%, 2nd: 93±16%, 3rd: 90±15%), regardless of the level of 1st visit restriction (Figure 2B, eTable 4). In contrast, patients with diffusion impairment at 1st visit remained, on average, diffusion impaired at all follow-up visits (DLCO 1st: 59±16%, 2nd: 66±19%, 3rd: 65±19%), with a clear association between worsening TLC and DLCO (Figure 2B, eTable 4). Additional PFT measurements are provided in eTable 4.

**Figure 2:**
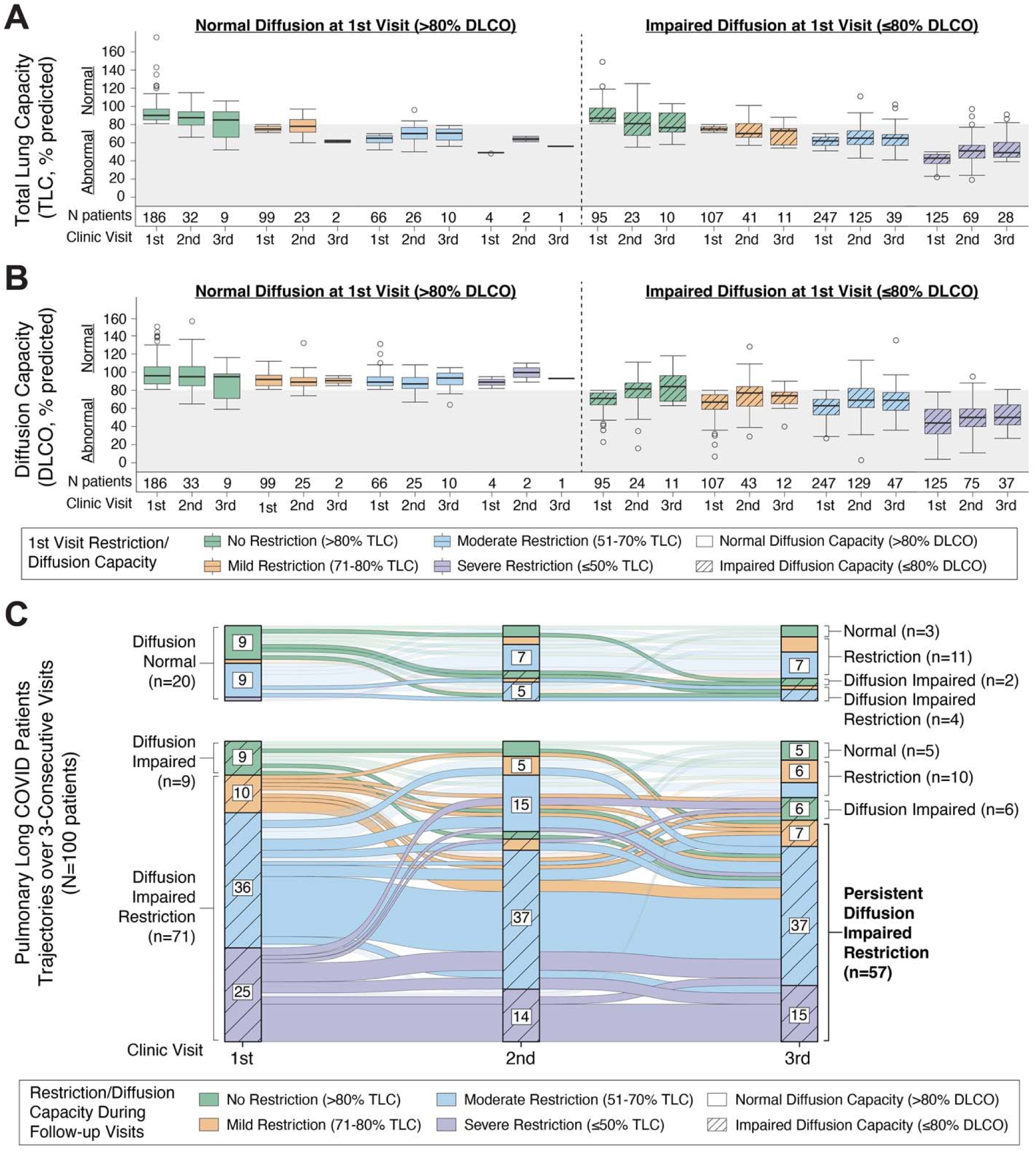
Diffusion Impaired Restriction is a Key Feature of Persistent Pulmonary Long COVID: (A-B) Results of percent predicted total lung capacity (TLC) and diffusing capacity for carbon monoxide (DLCO) are shown for 3 clinic visits with stratification by restriction severity (color) measured during the 1st visit PFT and presence or absence of diffusion impairment (hashed lines). Boxplots represent the median black center line; 25th and 75th percentile box boundaries) for each PFT measured, with number of patients (N) reported below each group. Normal TLC and DLCO are denoted by the grey color on the plot at 80%. (C) Alluvial diagram displays patient PFT trajectories over 3-consecutive visits (N=100 total) with relative improvement or decline as measured by TLC (color) and DLCO (hashed lines). Labelling of alluvial diagram axes groups with <5 patients was omitted for visual clarity.

Alluvial diagrams were used to assess improvement or decline in diffusion impairment and restriction for patients with 3-consecutive PFTs at the UAB pulmonary Long COVID clinic (N=100, Figure 2C). Overall, we observed that the majority of patients with diffusion impairment and severe or moderate restriction at 1st visit had persistent restriction and diffusion impairment at 2nd and 3rd follow-up visits (Figure 2C). Improvement from diffusion impairment at any level of restriction was rare, with only 5 patients regaining normal TLC and DLCO by their 3rd visit. Over half of the normal diffusion capacity patients (n=11) had some level of lung restriction at 1st visit (n=1 mild, n=9 moderate, n=1 severe). Restriction or diffusion impaired restriction were the predominant phenotypes observed by the 3rd follow-up visit, thereby indicating an earlier stage of disease followed by progression among the normal diffusion capacity patients (Figure 2C).

### Risk Factors for Developing Pulmonary Long COVID with Severe Pulmonary Restriction

Logistic regression models were used to identify risk factors for developing Long COVID with combined diffusion impairment (DLCO ≤80%) and severe or moderate restriction (TLC ≤70%). Unadjusted univariable modeling revealed advanced age ≥65 years (OR=1.63 [1.23-2.23]), male sex (OR=1.88 [1.43-2.48]), renal disease (OR=2.48 [1.474.15]), diabetes (OR=1.82 [1.30-2.51]), heart failure or hypertension (OR=2.54 [1.98-3.32]), smoking history (OR=1.39 [1.03-1.86]), ICU admission (OR=6.15 [4.24-9.36]), and use of nasal cannula (OR=4.21 [2.90-6.07]), high-flow nasal cannula (OR=6.10 [3.92-9.82]), and invasive mechanical ventilation (OR=16.0 [8.87-34.4]) as independent risk factors for developing pulmonary Long COVID with severe or moderate restriction (Table 2). After adjusting for all variables, we observed that invasive mechanical ventilation conferred the greatest increased odds of developing pulmonary Long COVID with diffusion impairment and severe or moderate restriction (aOR=10.9 [4.09-28.6]), followed by nasal cannula (aOR=4.0 [2.61-6.33]) and high-flow nasal cannula (aOR=3.86 [1.73-8.09]) use during primary SARS-CoV-2 infection, heart failure or hypertension (aOR=2.09 [1.46-2.99]) and male sex (aOR=1.44 [1.03-1.98]; Table 2; reference group: un-hospitalized primary infection patients). In a sub-analysis using only hospitalized patients (WHO score 4-7), the association of invasive mechanical ventilation with diffusion impairment and severe or moderate restriction was comparable with whole cohort results (aOR=9.56 [3.22-32.6] vs. reference group of hospitalized room-air patients; eTable 5). To evaluate the potential effect of patients with pre-existing pulmonary comorbidities on our model results, further sensitivity testing was performed (eTables 6-7). Overall, the results show that invasive mechanical ventilation still conferred the greatest risk for development of pulmonary Long COVID with diffusion impairment and severe or moderate restriction (eTable 6, cohort sans patients with pulmonary comorbidities vs. reference group of non-hospitalized patients aOR=14.5 [5.0-50.9]; eTable 7 cohort sans patients pulmonary comorbidities vs. reference group of hospitalized room-air patients aOR=15.2 [4.72-68.1]).

### Assessment of CT Imaging and Pathology

CT imaging was performed on a total of 308 patients (33%; n=246 diffusion impaired sub-group, n=62 diffusion normal sub-group; Table 1). CT pathology and increased lung involvement were predominantly found in diffusion impaired patients with severe or moderate restriction. CT scoring of images taken within 6-months of the 1st Long COVID visit identified ground glass opacities (85%), reticulations (82%), bronchiectasis (69%), and fibrotic changes (65%) as the defining pathologies in the majority of diffusion impaired severe restriction patients (Figure 3A, eTable 2). A similar pathologic profile was found in diffusion impaired patients with moderate restriction (Figure 3B). Univariable and multivariable logistic regression modeling was performed to determine if CT pathologies were associated with increased odds of developing pulmonary Long COVID with severe or moderate restriction (eTable 8). The significant unadjusted odds ratios for developing severe restriction were associated with ground glass opacities (OR=4.11 [2.50-6.96]), reticulations (OR=5.49 [3.24-9.35]), fibrotic changes (OR=5.27 [3.10-9.60]), bronchiectasis (OR=5.27 [3.10-9.60]), and consolidation (OR=2.35 [1.02-9.06]); however, only reticulations (aOR=2.12 [1.01-4.34]) maintained a significance in multivariable modeling (eTable 8).

**Figure 3:**
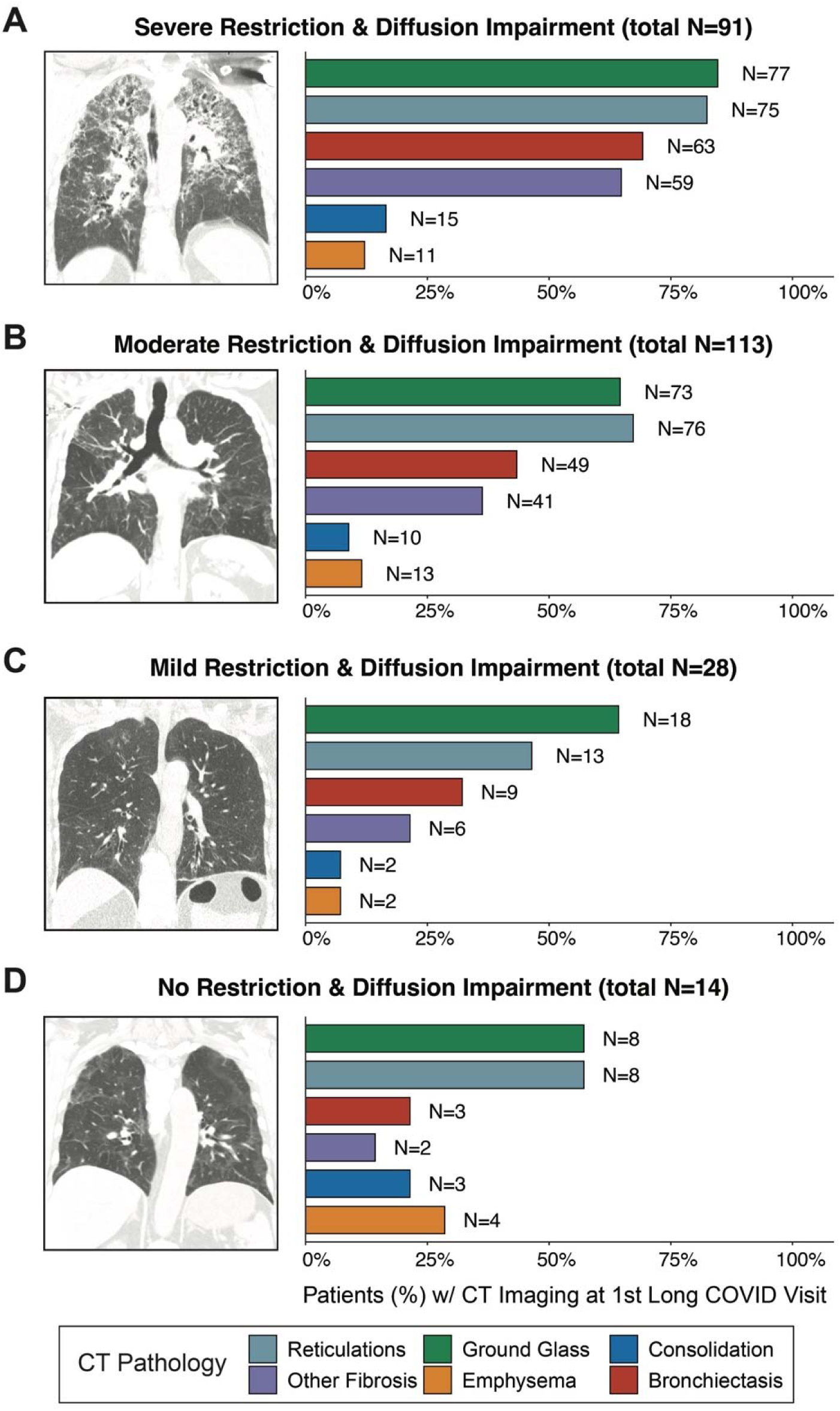
CT Image Findings in Pulmonary Long COVID with Diffusion Impaired Restriction: (A-D) Representative CT images of pulmonary Long COVID patients with diffusion impairment (DLCO ≤80%) with severe (TLC ≤50%), moderate (TLC 51-70%), mild (TLC 71-80%), and no restriction (TLC >80%) assessed at the 1st Long COVID clinic visit. Corresponding barcharts of CT pathology (%, N=patients) are displayed for each group. (Note: architectural distortion, traction bronchiectasis and honeycombing are termed ‘Other Fibrosis’).

## DISCUSSION

Our current understanding of persistent pulmonary defects from SARS-CoV-2 infection are primarily derived from prospective follow-up studies assessing patient outcomes after hospitalization with acute COVID-19. Gradual recovery of impaired diffusion capacity (DLCO ≤80%) and radiographic evidence of fibrotic pulmonary tissue are among the most commonly reported observations.^7–10,23–27^ Although these studies offer key insights into the trajectory of acute COVID recovery, characterizing patients without post-COVID symptoms limit the applicability of these findings to Long COVID patients with persistent dyspnea, cough, and chest discomfort. To address this knowledge gap, we leveraged a large, demographically diverse cohort comprised exclusively of Long COVID patients with persistent pulmonary symptoms. For this cohort, we aligned robust medical record data requiring no imputation, quantitative CT imaging, and longitudinal pulmonary function testing (PFT) to identify the characteristics and determinants of pulmonary Long COVID. Our study establishes a clear association between burden of acute COVID disease and the development of persistent lung restriction with diffusion impairment. Collectively, these results comprehensively define pathology for Long COVID that can be readily measured with PFTs and provides opportunity to better study the post-viral interplay of pulmonary symptoms with alterations in lung physiology.

Demographic risk factors (aOR [95% CI]) associated with the development of pulmonary Long COVID with diffusion impairment and severe or moderate restriction included male sex (aOR=1.44 [1.05-2.00]) and pre-existing heart failure and hypertension (aOR=2.07 [1.46-3.02]; Table 2, eTable 5). These findings differ from a recent meta-analysis of multi-organ Long COVID symptoms that noted an elevated risk in females (aOR=1.56) and individuals over 40 years of age (aOR=1.21).^28^ Our findings emphasize differences between the assessments obtained from broad symptom observations and physiological readouts in pinpointing at-risk populations. In Long COVID patients complaining of prolonged pulmonary symptoms, prior studies have suggested that dyspnea is a major feature and that symptoms can persist for months after initial infection.^9,29^ Our study affirmed this observation with 78% (n=725) of the UAB pulmonary Long COVID cohort identifying dyspnea as the primary symptom, followed by 18% (n=167) with cough and 4% (n=37) with chest discomfort (Table 1). Notably, the complaint of dyspnea remained consistent (>70%) across all degrees of diffusion impairment, restriction, and levels of lung involvement seen on CT imaging (Table 1, eTable 2). This suggests symptoms alone do not provide sufficient granularity to identify distinct endotypes of pulmonary Long COVID, thereby highlighting the importance of incorporating routine PFTs in the diagnostic evaluation of this patient population.

Our study demonstrates that the severity of hypoxia during primary SARS-CoV-2 infection is a critical factor in the development of pulmonary Long COVID with persistent diffusion impairment and restriction (Table 2). Notably, the post-hospitalization PFT impairments in pulmonary Long COVID contrast with previously described post ARDS findings in non-COVID patients, where subjects predominately present with isolated diffusion impairment which improves to normal levels over a 6 month-1 year period and limited lung restriction at any time-point.^30–33^ In addition to marked differences in lung physiology, the presence of reticulations, bronchiectasis, ground glass opacities, and fibrotic changes in pulmonary Long COVID CT images are distinct from ARDS which has been predominantly described by the presence of ground glass opacities and reticulations.^32^ Cumulatively, our physiologic and radiographic evidence suggest that pulmonary Long COVID is a pro-fibrotic disease process that is distinct from ARDS; however, future studies are warranted and needed to further elucidate the differences.

The biologic mechanisms underlying these symptomatic, physiologic, and radiographic changes are poorly understood, but there is increasing evidence that pro-fibrotic interstitial lung changes are occurring in dyspneic patients as early as 1 month post-COVID infection.^18,34–36^ From a molecular perspective, several independent lines of evidence have shown that altered immune function, dysregulation of systemic neutrophilic signatures, and persistent inflammation and presence of viral antigens are associated with Long COVID.^37–40^ While few studies have been conducted in the Long COVID lung, a prior spatial transcriptomic lung autopsy study from COVID-19 acute lung injury demonstrates a distinct fibro-proliferative phenotype relative to influenza infection.^41^ If true, pulmonary fibrosis therapeutics like Nintedanib and Pirfenidone, which have been sparingly used in post-COVID associated fibrosis, may be uniquely suited for the subset of pulmonary Long COVID patients with diffusion impaired restriction.^42,43,46^ This report provides novel definition of a distinct endotype of Long COVID and emphasizes the need to stratify patients for targeted therapeutic and clinical management.

This study has limitations. Due to the retrospective nature of the cohort, PFTs were not taken before or during primary SARS-CoV-2 infection, and therefore could not be compared to measurements taken at the first Long COVID clinic visit. While this large, demographically diverse cohort offers a unique opportunity to characterize pulmonary presentations of Long COVID, patients presented to the Long COVID clinic at different time intervals after primary COVID infection, and follow-ups were limited to attended visits. Similarly, inherent bias towards more severe disease is present in patients who had CT imaging performed. Despite these limitations, the combination of an objective physiologic metric in longitudinal PFTs and a previously established Long COVID symptomatic signature provide an important foundation for future Long COVID studies.

Although dyspnea is present in a majority of pulmonary Long COVID patients, reported symptoms were not representative of the degree of diffusion impairment and restriction across the cohort.^44^ The additional granularity provided with PFTs highlights the utility of a broadly available clinical test to identify pulmonary endotypes associated with persistent physiologic impairment. These insights underscore the need for medical providers to incorporate PFT measurements as a routine step for evaluating Long COVID patients with pulmonary complaints. Informed stratification of patients experiencing pulmonary Long COVID is critical as this population is likely to require high utilization of health care services and would likely benefit from early therapeutic interventions.^45^

## Data Availability

For protection of personal health information, data will only be made available with the approval of the University of Alabama at Birmingham IRB committee and the signing of formal data use agreement.

## AUTHOR CONTRIBUTIONS

Michael John Patton (MJP), Nathaniel Erdmann (NE), Donald Benson (DB), Scott Grumbley (SG), Emily B. Levitan (EBL), Amit Gaggar (AG), Sarah W. Robison (SWR), Raval Dhaval (RD), Kinner Patel (KP), Morgan L. Locy (MLL), Peter Morris (PM), and Matthew Might (MM) co-conceived the project, prepared the main text, and oversaw the UAB cohort creation. MJP processed and analyzed EMR data to create all tables and figures in the main text and supplement of this manuscript. DB analyzed and scored CT images. SG analyzed and scored a subset of CT images. We thank Dr. Jeanne Marrazzo, Dr. Surya Bhatt, and Dr. Mark Dransfield for contributing critical comments and additions to the main text and supplement. NE oversaw IRB approval from the University of Alabama at Birmingham (UAB IRBs: 300006291, 300006205). NE, MM, RD, and AG, obtained funding and supervised the overall study. All co-authors reviewed and approved the final version of the manuscript.

