## Supplemental Materials for "Characteristics and Determinants of Pulmonary Long COVID"

**Appendix**

| **eTable 1: Patient Characteristics Extracted from Electronic Medical Record (EMR) Data and CT Image Analysis used for Outcome Modeling** | | | |
| --- | --- | --- | --- |
| **Variable Type** | **Time Point** | **Variable Name** | **Variable Description** |
| Electronic  Medical Record  &  Chart Review | Before Primary  SARS-CoV-2  Infection | Diabetes (Type I or II) | Categorical (yes or no) |
|  |  | Renal Disease | Categorical (yes or no) |
|  |  | Pulmonary Disease | Categorical (yes or no) |
|  |  | Hypertension or Heart Failure | Categorical (yes or no) |
|  |  | Obstructive Sleep Apnea | Categorical (yes or no) |
|  |  | Vaccination Status (1^st^ dose) | Categorical (yes or no) |
|  |  | Smoking History | Categorical (current, former, or never) |
|  | During Primary  SARS-CoV-2  Infection | SARS-CoV-2 Infection Wave^a^ | Categorical  (Alpha, Delta, Omicron) |
|  |  | WHO Severity Score  (Worst score during entire hospitalization) | Categorical (3-7)   - (3) No Hospital Admission - (4) Admitted, Room Air - (5) Admitted, Nasal Cannula - (6) Admitted, High-Flow Nasal Cannula (>5L/min) - (7) Admitted, Invasive Mechanical Ventilation |
|  | 1^st^ Pulmonary  Long COVID  Clinic Visit | Age | Continuous (years) |
|  |  | Body-Mass Index | Continuous (kg/m^2^) |
|  |  | Biological Sex | Categorical  (male or female) |
|  |  | Time from primary infection to 1^st^ Long COVID clinic visit | Continuous (months) |
| CT Image  Analysis | Image taken within 6-months of  1^st^ Pulmonary Long COVID Clinic Visit | Right Upper Lung Involvement^b^ | Continuous (0-5) |
|  |  | Right Middle Lung Involvement^b^ | Continuous (0-5) |
|  |  | Right Lower Lung Involvement^b^ | Continuous (0-5) |
|  |  | Left Upper Lung  Involvement^b^ | Continuous (0-5) |
|  |  | Left Lower Lung Involvement^b^ | Continuous (0-5) |
|  |  | Cumulative Lung Involvement^b^ | Continuous (0-25) |
|  |  | Lung Consolidation | Categorical (yes or no) |
|  |  | Ground-Glass Opacities | Categorical (yes or no) |
|  |  | Reticulations | Categorical (yes or no) |
|  |  | Bronchiectasis | Categorical (yes or no) |
|  |  | Emphysema | Categorical (yes or no) |
|  |  | Other Fibrosis  (Architectural distortions, traction bronchiectasis, honeycombing) | Categorical (yes or no) |
| ^a^SARS-CoV-2 waves defined by following date ranges (Alpha: 01/01/2020-05/31/2021, Delta:06/01/2021-01/01/2022, Omicron: 01/02/2022-08/01/2023)  ^b^Lung Involvement was assessed with a 0-5 scale (0=no involvement, 1=1-5%, 2=5-25%, 3=25-50%, 4=50-75%, 5$\geq$75%) | | | |

| **eTable 2: Supplemental Cohort Characteristics of Pulmonary Long COVID Patients Stratified by Diffusion Capacity and Restriction in the UAB Cohort** | | | | | | | | | | | |
| --- | --- | --- | --- | --- | --- | --- | --- | --- | --- | --- | --- |
|  |  | | Diffusion Impaired (≤80% DLCO) | | | |  | Diffusion Normal (>80% DLCO) | | | |
|  | Restriction: | Severe | | Moderate | Mild | None | Restriction: | Severe | Moderate | Mild | None |
|  | TLC: | ≤50% | | 51-70% | 71-80% | >80% | TLC: | ≤50% | 51-70% | 71-80% | 80% |
|  | N=574 | n=125 | | n=247 | n=107 | n=95 | N=355 | n=4 | n=66 | n=99 | n=186 |
| Primary COVID Infection |  |  | |  |  |  |  |  |  |  |  |
| SARS-CoV-2  Variant Wave |  |  | |  |  |  |  |  |  |  |  |
| Alpha Wave | 357 (62) | 67 (54) | | 154 (62) | 69 (64) | 67 (71) | 225 (63) | 1 (25) | 48 (73) | 59 (60) | 117 (63) |
| Delta Wave | 130 (23) | 32 (26) | | 57 (23) | 25 (23) | 16 (17) | 74 (21) | 2 (50) | 10 (15) | 22 (22) | 40 (22) |
| Omicron Wave | 87 (15) | 26 (21) | | 36 (15) | 13 (12) | 12 (13) | 56 (16) | 1 (25) | 8 (12) | 18 (18) | 29 (16) |
| Immunosuppressed | 137 (38) | 38 (36) | | 68 (45) | 18 (30) | 13 (33) | 18 (19) | 0 (0) | 5 (22) | 4 (18) | 9 (18) |
| Unknown | 218 | 20 | | 97 | 46 | 55 | 260 | 3 | 43 | 77 | 137 |
| Remdesivir | 178 (46) | 60 (55) | | 78 (50) | 27 (42) | 13 (24) | 29 (19) | 1 (100) | 6 (17) | 6 (15) | 16 (21) |
| Unknown | 188 | 15 | | 90 | 42 | 41 | 200 | 3 | 31 | 58 | 108 |
| Dexamethasone | 239 (62) | 79 (72) | | 104 (65) | 37 (57) | 19 (35) | 45 (29) | 1 (100) | 14 (40) | 10 (24) | 20 (26) |
| Unknown | 186 | 15 | | 88 | 42 | 41 | 200 | 3 | 31 | 58 | 108 |
| COVID Severity (WHO Score) |  |  | |  |  |  |  |  |  |  |  |
| (3) No Admission | 217 (38) | 20 (16) | | 96 (39) | 46 (43) | 55 (58) | 260 (73) | 3 (75) | 43 (65) | 77 (78) | 137 (74) |
| (4) Room Air | 60 (10) | 8 (6) | | 21 (9) | 16 (15) | 15 (16) | 45 (13) | 0 (0) | 10 (15) | 11 (11) | 24 (13) |
| (5) Nasal Cannula | 139 (24) | 29 (23) | | 69 (28) | 24 (22) | 17 (18) | 32 (9) | 0 (0) | 8 (12) | 5 (5) | 19 (10) |
| (6) High-Flow Cannula | 88 (15) | 28 (22) | | 40 (16) | 15 (14) | 5 (5) | 15 (4) | 1 (25) | 3 (5) | 5 (5) | 6 (3) |
| (7) Ventilation | 70 (12) | 40 (32) | | 21 (9) | 6 (6) | 3 (3) | 3 (1) | 0 (0) | 2 (3) | 1 (1) | 0 (0) |
| Nasal Cannula Time (days)^1^ | 7 [4-11] | 8 [6-18] | | 7 [4-10] | 4 [4-7] | 7 [3-11] | 5 [3-6] | 6 [6-6] | 5 [3-6] | 5 [4-7] | 5 [4-6] |
| Unknown | 365 | 65 | | 147 | 78 | 75 | 318 | 3 | 57 | 92 | 166 |
| High-flow Nasal  Cannula Time (days)^1^ | 5 [3-8] | 8 [3-9] | | 4 [2-6] | 5 [2-9] | 4 [2-5] | 5 [2-9] | 9 [9-9] | 2 [2-2] | 11 [8-13] | 5 [3-9] |
| Unknown | 493 | 96 | | 213 | 95 | 89 | 345 | 3 | 64 | 97 | 181 |
| Ventilation Time (days)^1^ | 18 [8-35] | 27 [9-41] | | 13 [5-19] | 9 [7-11] | 24 [16-28] | 14 [10-19] | - | 14 [10-19] | - | - |
| Unknown | 516 | 90 | | 231 | 103 | 92 | 353 | 4 | 64 | 99 | 186 |
| CT Scoring Evaluations^2^ |  |  | |  |  |  |  |  |  |  |  |
| Cumulative  Involvement (0-25)^1^ | 9 [1-17] | 17 [9-21] | | 7 [1-13] | 3 [0-9] | 3 [1-7] | 0 [0-2] | 1 [1-1] | 0 [0-2] | 0 [0-2] | 0 [0-2] |
| Right Upper Lung (0-5) | 2 [0-3] | 3 [1-4] | | 1 [0-3] | 0 [0-2] | 0 [0-1] | 0 [0-0] | 0 [0-0] | 0 [0-0] | 0 [0-0] | 0 [0-0] |
| Right Middle Lung (0-5) | 2 [0-4] | 3 [2-5] | | 1 [0-3] | 0 [0-2] | 0 [0-1] | 0 [0-0] | 0 [0-0] | 0 [0-0] | 0 [0-0] | 0 [0-0] |
| Right Lower Lung (0-5) | 2 [0-4] | 3 [1-4] | | 1 [0-3] | 1 [0-2] | 1 [1-2] | 0 [0-1] | 0 [0-0] | 0 [0-1] | 0 [0-1] | 0 [0-1] |
| Left Upper Lung (0-5) | 2 [0-3] | 3 [2-4] | | 1 [0-2] | 0 [0-2] | 0 [0-1] | 0 [0-0] | 0 [0-0] | 0 [0-0] | 0 [0-1] | 0 [0-0] |
| Left Lower Lung (0-5) | 2 [1-4] | 3 [2-4] | | 1 [0-3] | 1 [0-2] | 1 [0-2] | 0 [0-1] | 1 [1-1] | 0 [0-1] | 0 [0-0] | 0 [0-1] |
| Unknown* | 328 | 34 | | 134 | 79 | 81 | 293 | 3 | 46 | 80 | 164 |
| CT Pathology |  |  | |  |  |  |  |  |  |  |  |
| Ground Glass Opacities | 176 (72) | 77 (85) | | 73 (65) | 18 (64) | 8 (57) | 16 (26) | 1 (100) | 4 (20) | 5 (26) | 6 (27) |
| Reticulations | 172 (70) | 75 (82) | | 76 (67) | 13 (46) | 8 (57) | 15 (24) | 0 (0) | 6 (30) | 5 (26) | 4 (18) |
| Other Fibrosis | 108 (44) | 59 (65) | | 41 (36) | 6 (21) | 2 (14) | 5 (8) | 0 (0) | 2 (10) | 1 (5) | 2 (9) |
| Bronchiectasis | 124 (50) | 63 (69) | | 49 (43) | 9 (32) | 3 (21) | 8 (13) | 0 (0) | 3 (15) | 2 (11) | 3 (14) |
| Consolidation | 30 (12) | 15 (16) | | 10 (9) | 2 (7) | 3 (21) | 1 (2) | 0 (0) | 0 (0) | 0 (0) | 1 (5) |
| Emphysema | 30 (12) | 11 (12) | | 13 (12) | 2 (7) | 4 (29) | 4 (6) | 0 (0) | 1 (5) | 2 (11) | 1 (5) |
| Unknown* | 328 | 34 | | 134 | 79 | 81 | 293 | 3 | 46 | 80 | 164 |
| Expiratory Scan  CT Pathology |  |  | |  |  |  |  |  |  |  |  |
| Gas Trapping  (Expiratory Only)^3^ | 118 (67) | 40 (57) | | 59 (74) | 16 (76) | 3 (60) | 24 (69) | 1 (100) | 8 (53) | 10 (83) | 5 (71) |
| Unknown | 398 | 55 | | 167 | 86 | 90 | 320 | 3 | 51 | 87 | 179 |
| ^1^Statistics are reported as n, (%) unless otherwise specified as Median [Q1-Q3]; N=total patients per diffusion capacity stratification group, n=total patients per restriction stratification group  ^2^Lung Involvement was assessed with a 0-5 scale (0=no involvement, 1=1-5%, 2=5-25%, 3=25-50%, 4=50-75%, 5$\geq$75%)  ^3^Gas trapping was evaluated in a small subset of patients with expiratory CT scans and was therefore not used as an outcome modeling variable.  *Unknown counts denoted with an asterix represent the count for all variables in the category (i.e. CT Scoring Evaluations, Median [IQR] and CT Pathology, n (%))  Abbreviations: TLC, percent predicted total lung capacity; DLCO, diffusion limitation of carbon monoxide; CT, computerized tomography;  WHO, World Health Organization; COVID, coronavirus disease; UAB, University of Alabama at Birmingham | | | | | | | | | | | |

| **eTable 3: CT Analysis Reader Agreement of Images taken within 6-months of the 1st Pulmonary Long COVID Clinic Visit** | | | |
| --- | --- | --- | --- |
|  | Reader #1,  N=91 | Reader #2,  N = 91 | p-value |
| CT Score - Right Upper Lung (0-5)^3^ |  |  | 0.68^1^ |
| 0 | 41 (45) | 36 (40) |  |
| 1 | 14 (15) | 21 (23) |  |
| 2 | 11 (12) | 11 (12) |  |
| 3 | 9 (10) | 7 (8) |  |
| 4 | 10 (11) | 7 (8) |  |
| 5 | 6 (7) | 9 (10) |  |
| CT Score - Right Middle Lung (0-5)^3^ |  |  | 0.38^1^ |
| 0 | 43 (47) | 36 (40) |  |
| 1 | 10 (11) | 20 (22) |  |
| 2 | 11 (12) | 10 (11) |  |
| 3 | 9 (10) | 7 (8) |  |
| 4 | 12 (13) | 9 (10) |  |
| 5 | 6 (7) | 9 (10) |  |
| CT Score - Right Lower Lung (0-5)^3^ |  |  | 0.96^1^ |
| 0 | 32 (35) | 32 (35) |  |
| 1 | 20 (22) | 21 (23) |  |
| 2 | 9 (10) | 9 (10) |  |
| 3 | 13 (14) | 9 (10) |  |
| 4 | 11 (12) | 12 (13) |  |
| 5 | 6 (7) | 8 (9) |  |
| CT Score - Left Upper Lung (0-5)^3^ |  |  | 0.043^1^ |
| 0 | 43 (47) | 37 (41) |  |
| 1 | 7 (8) | 20 (22) |  |
| 2 | 13 (14) | 11 (12) |  |
| 3 | 12 (13) | 7 (8) |  |
| 4 | 12 (13) | 7 (8) |  |
| 5 | 4 (4) | 9 (10) |  |
| CT Score - Left Lower Lung (0-5)^3^ |  |  | 0.35^1^ |
| 0 | 34 (37) | 30 (33) |  |
| 1 | 17 (19) | 21 (23) |  |
| 2 | 14 (15) | 7 (8) |  |
| 3 | 10 (11) | 14 (15) |  |
| 4 | 11 (12) | 9 (10) |  |
| 5 | 5 (5) | 10 (11) |  |
| CT Score - Cumulative Involvement^3^  (0-25), Median [Q1-Q3] | 5 [0-15] | 6 [1-15] | 0.60^2^ |
| Pathologies Present |  |  |  |
| Ground Glass Opacities, n (%) | 57 (63) | 66 (73) | 0.15^1^ |
| Reticulations, n (%) | 54 (59) | 37 (41) | 0.012^1^ |
| Other Fibrosis, n (%) | 28 (31) | 44 (48) | 0.015^1^ |
| Bronchiectasis, n (%) | 36 (40) | 40 (44) | 0.55^1^ |
| Consolidation, n (%) | 9 (10) | 12 (13) | 0.49^1^ |
| Emphysema, n (%) | 12 (13) | 12 (13) | >0.99^1^ |
| ^1^Pearson's Chi-squared test | | | |
| ^2^Wilcoxon rank sum test  ^3^Lung Involvement was assessed with a 0-5 scale (0=no involvement, 1=1-5%, 2=5-25%, 3=25-50%, 4=50-75%, 5$\geq$75%)  Statistics are reported as n (% of total) unless otherwise stated. | | | |

| **eTable 4: Longitudinal Pulmonary Function Testing Summary Statistics for 1st, 2nd, and 3rd, Pulmonary Long COVID Clinic Visits Stratified by 1st Visit Diffusion Impairment and Restriction** | | | | | | | | | | | | |
| --- | --- | --- | --- | --- | --- | --- | --- | --- | --- | --- | --- | --- |
|  |  | | Diffusion Impaired (≤80% DLCO) | | | |  | Diffusion Normal (>80% DLCO) | | | | |
| Percent Predicted PFT (%)^a^ | Restriction: | Severe | | Moderate | Mild | None | Restriction: | | Severe | Moderate | Mild | None |
|  | TLC: | ≤50% | | 51-70% | 71-80% | >80% | TLC: | | ≤50% | 51-70% | 71-80% | >80% |
| 1^st^ Long COVID  Clinic Visit^b^ | *N=574* | *n=125* | | *n=247* | *n=107* | *n=95* | *N=335* | | *n=4* | *n=66* | *n=99* | *n=186* |
| Total Lung Capacity (TLC) | 64[24] 65±18 | 43[10] 41±7 | | 62[9] 61±6 | 75[5] 75±3 | 87[15] 92±11 | 81[18] 82±15 | | 49[0] 49±1 | 65[8] 64±5 | 75[5] 75±3 | 90[12] 93±12 |
| Diffusion Capacity (DLCO) | 63[22] 59±16 | 44[27] 45±17 | | 63[17] 61±12 | 67[16] 65±14 | 71[13] 68±12 | 93[16] 95±12 | | 89[6] 89±6 | 89[10] 92±10 | 92[12] 92±8 | 96[19] 98±14 |
| Forced Vital Capacity (FVC) | 75[25] 74±18 | 52[15] 51±12 | | 74[17] 74±12 | 83[16] 84±12 | 91[18] 91±15 | 92[19] 93±14 | | 79[8] 76±9 | 82[11] 81±10 | 88[13] 88±10 | 99[17] 100±13 |
| Residual Volume  (RV) | 42[31] 47±31 | 26[18] 25±13 | | 39[23] 40±17 | 54[28] 56±21 | 74[47] 84±45 | 51[34] 55±31 | | 1[3] 3±3 | 36[22] 37±19 | 49[25] 47±19 | 62[35] 66±34 |
| Unknown | 6 | 5 | | 0 | 1 | 0 | 16 | | 1 | 8 | 7 | 0 |
| Forced Expiratory Volume 1-second (FEV1) | 75[25] 74±19 | 53[17] 55±13 | | 74[16] 74±15 | 85[17] 84±15 | 88[18] 87±17 | 91[18] 91±15 | | 77[12] 69±20 | 84[12] 83±10 | 87[16] 88±13 | 96[17] 97±15 |
| Unknown | - | - | | - | - | - | 1 | | 0 | 0 | 0 | 1 |
| Forced Mid-Expiratory Flow (FEF 25/75) | 83[47] 87±40 | 83[53] 86±45 | | 82[47] 86±38 | 90[48] 92±37 | 83[52] 88±39 | 90[39] 93±32 | | 71[26] 64±28 | 91[37] 94±27 | 88[44] 92±34 | 91[37] 94±32 |
| Unknown | 1 | 0 | | 0 | 0 | 1 | 2 | | 0 | 1 | 1 | 0 |
| 2^nd^ Long COVID  Clinic Visit^b^ | *N=282* | *n=77* | | *n=132* | *n=45* | *n=28* | *N=96* | | *n=3* | *n=27* | *n=29* | *n=37* |
| Total Lung Capacity (TLC) | 65[18] 65±15 | 51[14] 51±14 | | 65[15] 66±11 | 70[15] 73±10 | 81[25] 82±18 | 78[20] 79±13 | | 64[3] 64±4 | 70[13] 70±10 | 78[14] 79±10 | 88[15] 87±12 |
| Unknown | 24 | 8 | | 7 | 4 | 5 | 13 | | 1 | 1 | 6 | 5 |
| Diffusion Capacity (DLCO) | 67[28] 66±19 | 50[20] 51±16 | | 69[21] 70±16 | 77[22] 74±19 | 82[16] 76±20 | 89[16] 93±16 | | 100[11] 100±15 | 87[12] 88±11 | 89[9] 90±12 | 95[21] 98±20 |
| Unknown | 11 | 2 | | 3 | 2 | 4 | 11 | | 1 | 2 | 4 | 4 |
| Forced Vital Capacity (FVC) | 77[23] 76±17 | 59[16] 61±16 | | 80[19] 79±15 | 87[18] 85±12 | 88[26] 89±16 | 90[15] 90±14 | | 76[22] 74±22 | 86[10] 85±9 | 90[9] 90±11 | 96[19] 94±17 |
| Residual Volume (RV) | 41[28] 43±24 | 30[26] 33±22 | | 43[26] 44±22 | 44[29] 49±21 | 49[43] 56±33 | 55[33] 52±28 | | 33[30] 33±42 | 45[27] 49±27 | 47[31] 48±23 | 61[36] 57±30 |
| Unknown | 29 | 8 | | 12 | 4 | 5 | 16 | | 1 | 3 | 7 | 5 |
| Forced Expiratory Volume 1-second (FEV1) | 77[28] 76±19 | 61[18] 65±18 | | 80[26] 78±18 | 87[19] 85±14 | 86[19] 86±20 | 88[17] 88±16 | | 72[30] 65±30 | 86[13] 86±10 | 86[15] 88±11 | 91[20] 91±19 |
| Unknown | - | - | | - | - | - | 1 | | 0 | 0 | 0 | 1 |
| Forced Mid-Expiratory Flow (FEF 25/75) | 90[53] 91±42 | 93[63] 97±46 | | 86[50] 88±43 | 96[41] 94±32 | 89[53] 90±42 | 85[38] 89±31 | | 62[38] 54±39 | 95[42] 101±30 | 86[38] 87±22 | 77[35] 84±34 |
| 3^rd^ Long COVID  Clinic Visit^b^ | *N=114* | *n=39* | | *n=49* | *n=15* | *n=11* | *N=26* | | *n=1* | *n=10* | *n=2* | *n=13* |
| Total Lung Capacity (TLC) | 62[21] 64±15 | 49[16] 55±14 | | 65[12] 65±12 | 73[18] 69±12 | 77[21] 81±15 | 71[17] 72±15 | | 56[0] 56±NA | 71[12] 69±8 | 62[2] 62±2 | 85[28] 81±18 |
| Unknown | 26 | 11 | | 10 | 4 | 1 | 4 | | 0 | 0 | 0 | 4 |
| Diffusion Capacity (DLCO) | 66[25] 65±19 | 50[22] 54±14 | | 69[20] 68±18 | 74[13] 71±13 | 84[28] 84±18 | 94[18] 90±15 | | 93[0] 93±NA | 94[13] 91±13 | 91[6] 91±8 | 95[27] 88±20 |
| Unknown | 7 | 2 | | 2 | 3 | 0 | 4 | | 0 | 0 | 0 | 4 |
| Forced Vital Capacity (FVC) | 75[23] 75±17 | 61[18] 63±14 | | 79[16] 80±16 | 84[16] 83±11 | 86[17] 84±18 | 90[17] 89±16 | | 79[0] 79±NA | 91[9] 92±11 | 61[7] 61±10 | 94[19] 92±16 |
| Residual Volume (RV) | 37[27] 43±26 | 33[21] 36±22 | | 37[26] 42±25 | 41[47] 41±27 | 58[33] 64±36 | 41[27] 42±20 | | 27[0] 27±NA | 33[19] 33±17 | 46[28] 46±39 | 53[25] 54±15 |
| Unknown | 28 | 11 | | 11 | 5 | 1 | 5 | | 0 | 0 | 0 | 5 |
| Forced Expiratory Volume 1-second (FEV1) | 78[26] 77±20 | 65[19] 67±17 | | 80[20] 82±20 | 87[17] 85±11 | 82[36] 80±24 | 91[19] 88±18 | | 79[0] 79±NA | 93[16] 92±15 | 61[15] 61±21 | 91[13] 90±18 |
| Forced Mid-Expiratory Flow (FEF 25/75) | 98[61] 100±52 | 112[68] 103±52 | | 95[62] 100±58 | 96[29] 101±29 | 91[63] 86±49 | 90[46] 93±37 | | 79[0] 79±NA | 95[32] 98±35 | 81[54] 81±76 | 90[47] 91±37 |
| ^a^Percent Predicted Pulmonary Function Testing (PFT) values are represented as median [IQR], mean ± standard deviation.  ^b^N represents the overall patient count; n represents the stratified group patient count | | | | | | | | | | | | |

| **eTable 5: Outcome Modeling of Pulmonary Long COVID with Diffusion Impaired Restriction in Hospitalized COVID Patients (Sensitivity Testing)** | | | | |
| --- | --- | --- | --- | --- |
|  | Patients Total N | Diffusion Impaired Restriction^4^ N (% of Total) | Diffusion Impaired Restriction Unadjusted OR [95% CI]^1,2^ | Diffusion Impaired Restriction Adjusted OR [95% CI]^2,3^ |
| Advanced Age |  |  |  |  |
| <65 years | 342 | 188 (55) | — | — |
| ≥65 years | 110 | 68 (62) | 1.33 [0.85-2.06] | 1.05 [0.60-1.84] |
| Sex |  |  |  |  |
| Female | 278 | 138 (50) | — | — |
| Male | 174 | 118 (68) | 2.08 [1.46-3.22] | 1.82 [1.14-2.87] |
| Elevated BMI |  |  |  |  |
| <30 | 160 | 90 (56) | — | — |
| ≥30 | 292 | 166 (57) | 1.02 [0.69-1.54] | 1.17 [0.71-1.91] |
| Pulmonary Disease |  |  |  |  |
| No | 338 | 191 (57) | — | — |
| Yes | 114 | 65 (57) | 1.01 [0.65-1.57] | 1.14 [0.69-1.99] |
| Renal Disease |  |  |  |  |
| No | 398 | 220 (55) | — | — |
| Yes | 54 | 36 (67) | 1.64 [0.92-3.12] | 1.01 [0.51-2.45] |
| Diabetes |  |  |  |  |
| No | 344 | 189 (55) | — | — |
| Yes | 108 | 67 (62) | 1.33 [0.86-2.13] | 1.21 [0.68-2.23] |
| Heart Failure or Hypertension |  |  |  |  |
| No | 203 | 96 (47) | — | — |
| Yes | 249 | 160 (64) | 2.00 [1.40-2.91] | 2.12 [1.28-3.74] |
| Obstructive Sleep Apnea |  |  |  |  |
| No | 338 | 197 (58) | — | — |
| Yes | 114 | 59 (52) | 0.76 [0.52-1.21] | 0.56 [0.33-0.95] |
| Smoking History |  |  |  |  |
| Never smoker | 303 | 166 (55) | — | — |
| Current or Former  Smoker | 149 | 90 (60) | 1.25 [0.85-1.92] | 1.04 [0.62-1.74] |
| Vaccination Status |  |  |  |  |
| No | 386 | 217 (56) | — | — |
| Yes | 66 | 39 (59) | 1.13 [0.67-1.99] | 1.65 [0.77-3.39] |
| Months from Primary Infection to Long COVID Clinic Visit |  |  |  |  |
| 1-3 Months | 156 | 91 (58) | — | — |
| 3-6 Months | 165 | 109 (66) | 1.38 [0.91-2.21] | 1.20 [0.70-2.08] |
| 6-12 Months | 73 | 30 (41) | 0.51 [0.27-0.89] | 0.56 [0.28-1.11] |
| >12 Months | 58 | 26 (45) | 0.58 [0.32-1.12] | 0.53 [0.25-1.14] |
| ICU Admission |  |  |  |  |
| No | 291 | 134 (46) | — | — |
| Yes | 161 | 122 (76) | 3.64 [2.44-5.77] | 1.56 [0.72-3.54] |
| COVID Severity  (WHO Score) |  |  |  |  |
| (4) Room Air | 105 | 29 (28) | — | — |
| (5) Nasal Cannula | 171 | 98 (57) | 3.53 [2.11-6.30] | 3.84 [2.14-7.62] |
| (6) High-Flow Cannula | 103 | 68 (66) | 5.21 [2.78-9.76] | 3.76 [1.59-10.8] |
| (7) Ventilation | 73 | 61 (84) | 13.8 [6.78-35.1] | 9.56 [3.22-32.6] |
| ^1^Unadjusted Odds Ratio 95% confidence interval (n=1000 bootstraps) | | | | |
| ^2^OR = Odds Ratio, CI = Confidence Interval | | | | |
| ^3^Adjusted Odds Ratio 95% confidence interval (n=1000 bootstraps)  ^4^Diffusion impaired restriction is defined by a DLCO ≤80% and a TLC ≤70% measured by PFT at the 1^st^ Long COVID clinic visit | | | | |

| **eTable 6:Outcome Modeling of Pulmonary Long COVID with Diffusion Impaired Restriction in Patients without Pre-Existing Pulmonary Comorbidities**  **(Sensitivity Testing)** | | | | |
| --- | --- | --- | --- | --- |
|  | Patients Total N | Diffusion  Impaired Restriction^4^ N (% of Total) | Diffusion  Impaired Restriction Unadjusted OR [95% CI]^1,2^ | Diffusion  Impaired Restriction Adjusted OR [95% CI]^2,3^ |
| Advanced Age |  |  |  |  |
| <65 years | 581 | 218 (38) | — | — |
| ≥65 years | 137 | 68 (50) | 1.65 [1.10-2.48] | 1.27 [0.79-2.04] |
| Sex |  |  |  |  |
| Female | 463 | 161 (35) | — | — |
| Male | 255 | 125 (49) | 1.79 [1.34-2.45] | 1.39 [0.97-2.03] |
| Elevated BMI |  |  |  |  |
| <30 | 292 | 107 (37) | — | — |
| ≥30 | 426 | 179 (42) | 1.26 [0.95-1.72] | 1.38 [0.96-1.99] |
| Renal Disease |  |  |  |  |
| No | 663 | 251 (38) | — | — |
| Yes | 55 | 35 (64) | 2.87 [1.69-5.25] | 1.26 [0.61-2.69] |
| Diabetes |  |  |  |  |
| No | 582 | 216 (37) | — | — |
| Yes | 136 | 70 (51) | 1.82 [1.23-2.72] | 1.14 [0.71-1.89] |
| Heart Failure or Hypertension |  |  |  |  |
| No | 402 | 122 (30) | — | — |
| Yes | 316 | 164 (52) | 2.47 [1.82-3.47] | 2.08 [1.39-3.18] |
| Obstructive Sleep Apnea |  |  |  |  |
| No | 567 | 233 (41) | — | — |
| Yes | 151 | 53 (35) | 0.76 [0.51-1.10] | 0.42 [0.25-0.67] |
| Smoking History |  |  |  |  |
| Never smoker | 507 | 189 (37) | — | — |
| Current or Former  Smoker | 211 | 97 (46) | 1.43 [1.00-2.00] | 1.38 [0.92-2.00] |
| Vaccination Status |  |  |  |  |
| No | 585 | 237 (41) | — | — |
| Yes | 133 | 49 (37) | 0.86 [0.57-1.24] | 1.11 [0.68-1.76] |
| Months from Primary Infection to Long COVID Clinic Visit |  |  |  |  |
| 1-3 Months | 232 | 101 (44) | — | — |
| 3-6 Months | 227 | 106 (47) | 1.13 [0.78-1.66] | 0.89 [0.57-1.37] |
| 6-12 Months | 147 | 43 (29) | 0.53 [0.33-0.83] | 0.66 [0.40-1.12] |
| >12 Months | 112 | 36 (32) | 0.61 [0.38-1.01] | 0.66 [0.37-1.18] |
| ICU Admission |  |  |  |  |
| No | 595 | 196 (33) | — | — |
| Yes | 123 | 90 (73) | 5.62 [3.66-8.81] | 1.15 [0.51-2.55] |
| COVID-19 Severity |  |  |  |  |
| (3) No Admission | 380 | 95 (25) | — | — |
| (4) Room Air | 72 | 20 (28) | 1.17 [0.63-2.02] | 0.96 [0.53-1.78] |
| (5) Nasal Cannula | 130 | 72 (55) | 3.78 [2.50-5.70] | 3.47 [2.23-5.75] |
| (6) High-Flow  Cannula | 83 | 54 (65) | 5.67 [3.33-9.58] | 4.09 [1.87-9.54] |
| (7) Ventilation | 53 | 45 (85) | 17.2 [8.95-45.6] | 14.5 [5.00-50.9] |
| ^1^Unadjusted Odds Ratio 95% confidence interval (n=1000 bootstraps) | | | | |
| ^2^OR = Odds Ratio, CI = Confidence Interval | | | | |
| ^3^Adjusted Odds Ratio 95% confidence interval (n=1000 bootstraps)  ^4^Diffusion impaired restriction is defined by a DLCO ≤80% and a TLC ≤70% measured by PFT at the 1^st^ Long COVID clinic visit | | | | |

| **eTable 7: Outcome Modeling of Pulmonary Long COVID with Diffusion Impaired Restriction in Hospitalized COVID Patients without Pre-Existing Pulmonary Comorbidities (Sensitivity Testing)** | | | | |
| --- | --- | --- | --- | --- |
|  | Patients Total N | Diffusion  Impaired Restriction^4^ N (% of Total) | Diffusion  Impaired Restriction Unadjusted OR [95% CI]^1,2^ | Diffusion  Impaired Restriction Adjusted OR [95% CI]^2,3^ |
| Advanced Age |  |  |  |  |
| <65 years | 259 | 141 (54) | — | — |
| ≥65 years | 79 | 50 (63) | 1.44 [0.84-2.46] | 1.06 [0.53-2.11] |
| Sex |  |  |  |  |
| Female | 205 | 102 (50) | — | — |
| Male | 133 | 89 (67) | 2.06 [1.31-3.24] | 1.67 [0.96-3.01] |
| Elevated BMI |  |  |  |  |
| <30 | 126 | 70 (56) | — | — |
| ≥30 | 212 | 121 (57) | 1.06 [0.65-1.66] | 1.43 [0.78-2.71] |
| Renal Disease |  |  |  |  |
| No | 292 | 159 (54) | — | — |
| Yes | 46 | 32 (70) | 1.96 [1.00-4.20] | 1.21 [0.44-3.06] |
| Diabetes |  |  |  |  |
| No | 249 | 135 (54) | — | — |
| Yes | 89 | 56 (63) | 1.45 [0.88-2.42] | 1.31 [0.65-2.77] |
| Heart Failure or Hypertension |  |  |  |  |
| No | 152 | 71 (47) | — | — |
| Yes | 186 | 120 (65) | 2.11 [1.33-3.25] | 2.31 [1.21-4.80] |
| Obstructive Sleep Apnea |  |  |  |  |
| No | 255 | 151 (59) | — | — |
| Yes | 83 | 40 (48) | 0.62 [0.36-1.05] | 0.39 [0.21-0.76] |
| Smoking History |  |  |  |  |
| Never smoker | 235 | 129 (55) | — | — |
| Current or Former  Smoker | 103 | 62 (60) | 1.25 [0.79-2.02] | 1.10 [0.57-2.10] |
| Vaccination Status |  |  |  |  |
| No | 292 | 165 (57) | — | — |
| Yes | 46 | 26 (57) | 1.02 [0.53-1.97] | 1.58 [0.65-3.73] |
| Months from Primary Infection to Long  COVID Clinic Visit |  |  |  |  |
| 1-3 Months | 114 | 67 (59) | — | — |
| 3-6 Months | 124 | 80 (65) | 1.27 [0.72-2.23] | 1.07 [0.54-2.02] |
| 6-12 Months | 50 | 22 (44) | 0.55 [0.26-1.06] | 0.61 [0.25-1.47] |
| >12 Months | 50 | 22 (44) | 0.56 [0.27-1.08] | 0.58 [0.22-1.36] |
| ICU Admission |  |  |  |  |
| No | 217 | 101 (47) | — | — |
| Yes | 121 | 90 (74) | 3.35 [2.15-5.43] | 1.20 [0.53-3.09] |
| COVID-19 Severity |  |  |  |  |
| (4) Room Air | 72 | 20 (28) | — | — |
| (5) Nasal Cannula | 130 | 72 (55) | 3.35 [1.80-6.30] | 3.86 [1.79-9.17] |
| (6) High-Flow Cannula | 83 | 54 (65) | 5.04 [2.46-10.0] | 4.72 [1.78-15.9] |
| (7) Ventilation | 53 | 45 (85) | 15.1 [6.47-46.8] | 15.2 [4.72-68.1] |
| ^1^Unadjusted Odds Ratio 95% confidence interval (n=1000 bootstraps) | | | | |
| ^2^OR = Odds Ratio, CI = Confidence Interval | | | | |
| ^3^Adjusted Odds Ratio 95% confidence interval (n=1000 bootstraps)  ^4^Diffusion impaired restriction is defined by a DLCO ≤80% and a TLC ≤70% measured by PFT at the 1^st^ Long COVID clinic visit | | | | |

| **eTable 8: Outcome Modeling of Pulmonary Long COVID with Diffusion Impaired Restriction from CT Image Pathology** | | | | |
| --- | --- | --- | --- | --- |
|  | Patients Total N | Diffusion  Impaired Restriction^4^ N (% of Total) | Diffusion  Impaired Restriction Unadjusted OR [95% CI]^1,2^ | Diffusion Impaired Restriction Adjusted OR [95% CI]^2,3^ |
| Ground Glass Opacities |  |  |  |  |
| No | 116 | 54 (47) | — | — |
| Yes | 192 | 150 (78) | 4.11 [2.50-6.96] | 1.40 [0.75-2.78] |
| Reticulations |  |  |  |  |
| No | 121 | 53 (44) | — | — |
| Yes | 187 | 151 (81) | 5.49 [3.24-9.35] | 2.12 [1.01-4.34] |
| Other Fibrosis |  |  |  |  |
| No | 195 | 104 (53) | — | — |
| Yes | 113 | 100 (88) | 6.79 [3.59-15.0] | 2.30 [0.95-5.96] |
| Bronchiectasis |  |  |  |  |
| No | 176 | 92 (52) | — | — |
| Yes | 132 | 112 (85) | 5.27 [3.10-9.60] | 2.00 [0.92-4.82] |
| Consolidation |  |  |  |  |
| No | 277 | 179 (65) | — | — |
| Yes | 31 | 25 (81) | 2.35 [1.02-9.06] | 1.39 [0.57-5.40] |
| Emphysema |  |  |  |  |
| No | 274 | 180 (66) | — | — |
| Yes | 34 | 24 (71) | 1.27 [0.60-3.15] | 1.06 [0.42-2.99] |
| ^1^Unadjusted Odds Ratio 95% confidence interval (n=1000 bootstraps) | | | | |
| ^2^OR = Odds Ratio, CI = Confidence Interval | | | | |
| ^3^Adjusted Odds Ratio 95% confidence interval (n=1000 bootstraps)  ^4^Diffusion impaired restriction is defined by a DLCO ≤80% and a TLC ≤70% measured by PFT at the 1^st^ Long COVID clinic visit | | | | |
